# Clinical validation of a statin-benefit polygenic score using real-world cohorts of primary prevention participants

**DOI:** 10.1101/2025.10.09.25337698

**Authors:** Tanushree Haldar, Kyung Min Lee, Craig C. Teerlink, Kruthika R. Iyer, Amy L. Voorhees, James A. Najera, Nanase Toda, Meng Lu, Michael P. Douglas, Alice B. Popejoy, Adam Bress, Catherine Tcheandjieu, Philip S. Tsao, Kyong-Mi Chang, Ronald M. Krauss, Neil Risch, Carlos Iribarren, Julie A. Lynch, Akinyemi Oni-Orisan

**Affiliations:** Department of Clinical Pharmacy, University of California San Francisco, San Francisco, CA, USA; VA Informatics and Computing Infrastructure (VINCI), VA Salt Lake City Health Care System, Salt Lake City, UT, USA; Division of Epidemiology, Department of Internal Medicine, University of Utah School of Medicine, Salt Lake City, UT, USA; Department of Medicine, Stanford University School of Medicine, Stanford, CA, USA; Palo Alto VA Healthcare System, Palo Alto, CA, USA; Gladstone Institute of Data Science and Biotechnology, Gladstone Institutes, San Francisco, CA, USA; Department of Epidemiology and Biostatistics, University of California San Francisco, San Francisco, CA, USA; Kaiser Permanente Northern California Division of Research, Pleasanton, CA, USA; Department of Public Health Sciences (Epidemiology Division), School of Medicine, University of California, Davis, Davis, CA, USA; University of California Davis Comprehensive Cancer Center, University of California Davis Health, Sacramento, CA, USA; Department of Population Health Sciences, Division of Health System Innovation and Research, University of Utah School of Medicine, Salt Lake City, UT, USA; Institute for Human Genetics, University of California San Francisco, San Francisco, CA, USA; Department of Medicine, University of Pennsylvania Perelman School of Medicine, Philadelphia, PA, USA; Department of Pediatrics, University of California San Francisco, San Francisco, CA, USA; Department of Medicine, University of California San Francisco, San Francisco, CA, USA

## Abstract

**Background:** Polygenic risk scores derived from coronary artery disease genome-wide association studies are associated with statin relative risk reduction.

**Objective:** Examine the relationship between coronary artery disease polygenic risk scores that include variants below thresholds of genome-wide significance and statin primary prevention of major adverse cardiovascular events.

**Methods:** We generated coronary artery disease polygenic risk scores for participants with no past evidence of myocardial infarction from three electronic health record-linked genetic biobanks: All of Us Research Program, Genetic Epidemiology Research on Adult Health and Aging, and the Million Veteran Program. We scored each participant using three different polygenic risk scores: two with both genome-wide and sub-genome-wide significant variants (metaGRS and PRS2022) and one with only variants meeting genome-wide significance (164SNP). We used covariate-adjusted Cox regression models to compare risk of major adverse cardiovascular events between statin users and nonusers matched on age, sex, smoking status, type 2 diabetes mellitus, and hypertension within strata defined by polygenic risk. For the primary analysis, we performed a meta-analysis across the three cohorts for the delta hazard ratio of statin effectiveness between high and low polygenic risk.

**Results:** Across all cohorts, statin use was more strongly associated with reduced risk of major adverse cardiovascular events among participants with no myocardial infarction at index in the highest versus lowest polygenic risk score group for the PRS2022 (interaction beta 0.19, standard error 0.07, interaction *P*=2.3E-3) and metaGRS (interaction beta 0.14, standard error 0.07, interaction *P*=.02) scores. However, the association was not statistically significant (interaction beta 0.09, standard error 0.07, interaction *P*=.08) for the 164SNP risk score.

**Conclusions:** We demonstrated that the association between coronary artery disease polygenic risk scores and statin relative risk reduction can by enhanced with the inclusion of sub-genome-wide variants, paving the way for more research to establish clinical utility.

## Introduction

Improved optimization of statin prescribing has the potential to prevent adverse atherosclerotic cardiovascular disease (ASCVD) events among the greater than 75 million people receiving or guideline-eligible for therapy in the United States.^1^ However, traditional risk factors for ASCVD do not modify statin efficacy independent of statin-induced low-density lipoprotein cholesterol (LDL-C) lowering.^2^ In prior substudies using statin randomized controlled trial data post-hoc, polygenic risk scores derived from coronary artery disease genome-wide association studies were strongly associated with statin-relative risk reduction independent of LDL-C lowering and other clinical risk factors for ASCVD.^3,4^ Similar results were recently observed in real-world participants, suggesting that statin polygenic scores predict therapeutic response beyond idealistic clinical trial settings.^5^ However, findings from that initial study were generated in a single cohort of participants that may not represent statin users across the United States broadly. Furthermore, only one type of coronary artery disease polygenic risk score had been investigated in the context of statin relative risk reduction. To better determine the generalizability of these results, we investigated the relationship between polygenic score and statin coronary artery disease therapy effectiveness in multiple large electronic health record (EHR)-linked biobanks using various scoring types.

## Methods

### Data source

We conducted our studies using data from three independent EHR-linked biobanks including: the All of Us Research Program (AoURP), Genetic Epidemiology Research on Adult Health and Aging (GERA), and the Million Veteran Program (MVP). Regulatory approval for these secondary data analyses were obtained from Kaiser Permanente Mid-Atlantic States, University of California San Francisco, and Veterans Health Administration (VHA) Central Institutional Review Boards. Further details on the data sources are available in the Supplementary Methods. Diagnostic/procedure codes used in this study are available in Supplementary Table 1.

### Study population

To obtain a primary prevention population, we included participants with no evidence of myocardial infarction prior to index. This definition for primary prevention was based on published studies that also investigated the association between coronary artery disease polygenic risk scores and statin ASCVD benefit.^3–5^ Among these participants, we also used a smaller subset of participants without evidence of any ASCVD prior to the time of index. For this purpose, we defined ASCVD as evidence of coronary artery disease (myocardial infarction, coronary artery bypass graft, coronary revascularization, unstable angina, and stable angina), peripheral artery disease, cerebrovascular disease (ischemic stroke, transient ischemia attack, carotid stenosis), or abdominal aortic aneurysm.^6^ The development of ASCVD following index date was not a criterion for exclusion. See Supplementary Table 1 for further details on the primary prevention populations.

### Polygenic scores

We selected three different polygenic risk scores of coronary artery disease to be used as our statin ASCVD benefit polygenic scores.^5,7,8^ Two scores (metaGRS and PRS2022) each included both genome-wide and sub-genome-wide significant variants.^7,8^ The third score (164SNP) included only variants meeting genome-wide significance.^5^

Polygenic score for each participant was determined using PLINK2. Participants were grouped together within each of the three scores, as previously described.^5^ Briefly, participants were ranked by increasing score and divided into quintiles. Participants were organized into low, intermediate, and high polygenic score groups based on quintiles 1, 2 through 4, and 5 respectively. These polygenic risks scores, similar to most, were originally derived from the genome-wide association studies of Eurocentrically biased study populations and have not shown strong associations in other groups for statin benefit.^3–5^ Thus, after careful consideration, we used participants with genetic background most similar to the “EUR” superpopulation of the 1000 Genomes Project (i.e., 1KG-EUR-like) for this validation study. We also consulted recent guidance on the use of population descriptors in genomics research.^9^

### Drug response phenotype

We used a similar statin effectiveness phenotype as previously described.^5^ Briefly, a participant met the definition of a statin user if (1) they were considered to be adherent from the date of statin initiation to the date of event or censor; (2) they appeared to be a new initiator of statin therapy within the time frame of follow-up; (3) their last statin dispensing record window ended ≤30 days before the date of event or censor; (4) their date of statin initiation occurred >30 days before the date of event or censor (time-to-event and censoring analyses described in more detail below); and (5) had ≥1 complete lipid panel measurement prior to statin initiation. For MVP, >90 days of cumulative exposure was also required for users. For AoURP, participants also required a ≥25% lowering of LDL-C to be deemed a user.

A participant met the definition of a statin nonuser if they had no statin dispensing records before the date of event or censoring unless statin initiation was considered too recent to impact outcomes (i.e., ≤30 days before event or censoring).

To identify statin nonusers with similar ASCVD risk as statin users at index, we matched each user to two nonusers by age (within 3 years at a maximum), sex, cigarette smoking status, type 2 diabetes mellitus, hypertension, and self-identified race/ethnicity. For statin nonusers, the date of a lipid panel measurement that minimized age difference between a user and matched nonusers was set as the index timepoint. We also calculated the 10-year risk from index for a first-time hard ASCVD event in statin users and nonusers using the Pooled Cohort Equations (which use age, sex, smoking, diabetes, self-identified race, systolic blood pressure, use of antihypertensive therapy, total cholesterol, and high-density lipoprotein cholesterol to determine risk) to confirm similar risk between groups.^10^ See the Supplementary Methods for further details on our approach with the Pooled Cohort Equations.

Statin effectiveness was determined from hazard models of statin users and nonusers using time-to-first-event (from index) data with right-censoring for the outcomes described below. Participants who did not experience an outcome were censored at the time of death or the last LDL-C measurement in their records, whichever occurred first (GERA participants were also censored at age 90, and AoURP participants were also censored at the time of the last body mass index). Cox proportional hazards regression models used to generate the hazard ratios (HR) included sex, age, hypertension, type 2 diabetes mellitus, and cigarette smoking status at the time of index date as covariates.

### Outcomes

Consistent with the majority of statin randomized controlled trials, we used major adverse cardiovascular events as our cardiovascular outcome of interest. Major adverse cardiovascular events were defined as any of the following: myocardial infarction, coronary revascularization, unstable angina, ischemic stroke, transient ischemia attack, or death. See Supplementary Table 1 for further details on this outcome.

### Statistical analyses

Based on findings from a prior study,^5^ our prespecified primary analysis was the meta-analysis across all 3 cohorts for the delta HR of statin effectiveness between high and low polygenic score groups (interaction P-value) from each of the 3 coronary artery disease polygenic risk scores. The secondary analysis investigated the relationship between polygenic score and time-to-first-event in statin nonusers only. All analyses were conducted using R software (R Foundation for Statistical Computing, version 3.5.1, https://www.R-project.org/; Vienna, Austria).

## Results

### Study population

The study population was made up of 50,949 total participants across the 3 cohorts (16,983 statin users). The characteristics of each cohort varied at index. For example, 92% of the population were assigned male at birth in MVP compared to 45% and 51% for GERA and AoURP, respectively. Table 1 provides further details for each cohort.

**Table 1.**
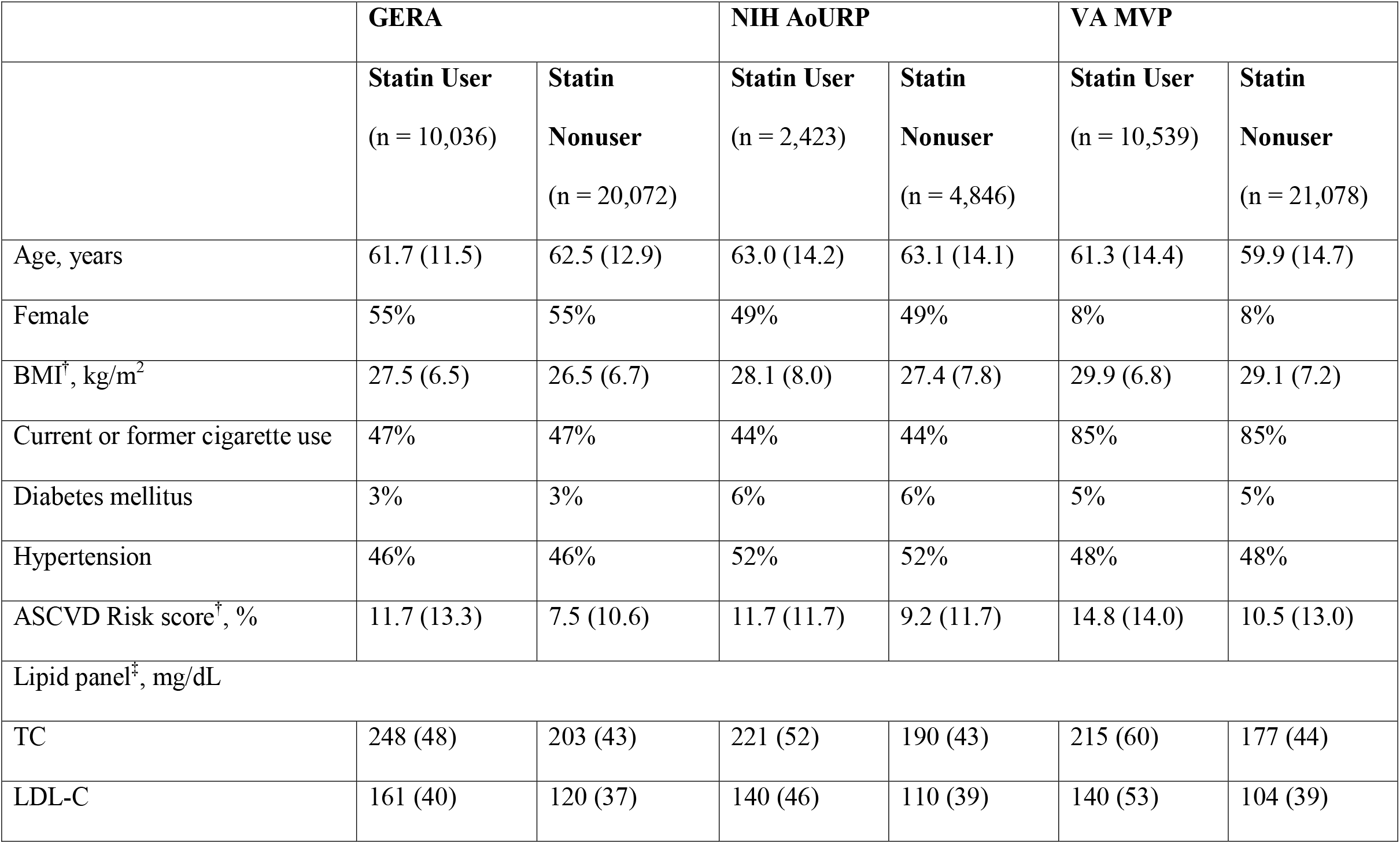
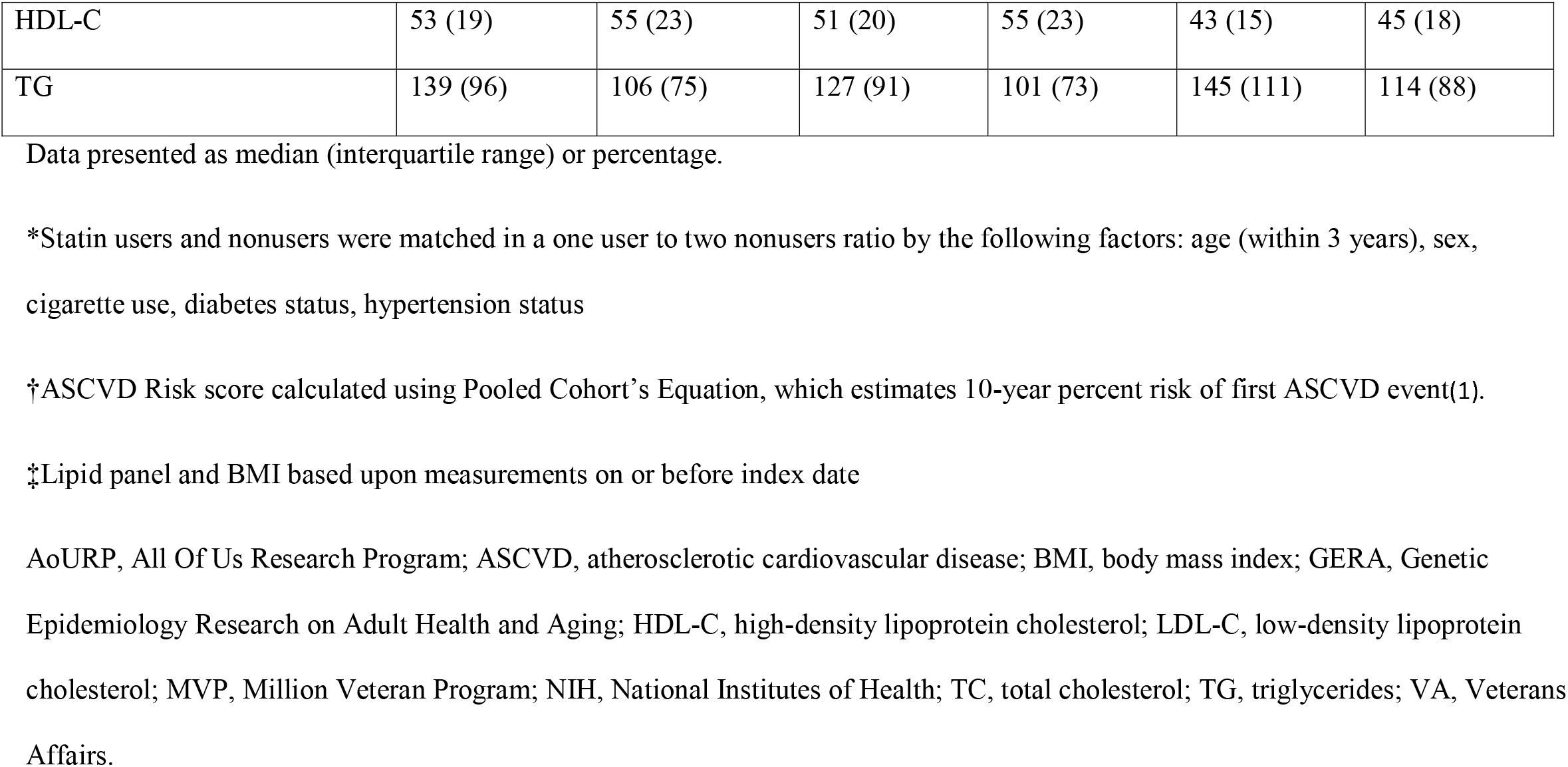
Clinical characteristics of statin users and matched nonusers* at index.

|  | **GERA** |  | **NIH AoURP** |  | **VA MVP** |  |
| --- | --- | --- | --- | --- | --- | --- |
|  | **Statin User**<br>(n = 10,036) | **Statin Nonuser**<br>(n = 20,072) | **Statin User**<br>(n = 2,423) | **Statin Nonuser**<br>(n = 4,846) | **Statin User**<br>(n = 10,539) | **Statin Nonuser**<br>(n = 21,078) |
| Age, years | 61.7 (11.5) | 62.5 (12.9) | 63.0 (14.2) | 63.1 (14.1) | 61.3 (14.4) | 59.9 (14.7) |
| Female | 55% | 55% | 49% | 49% | 8% | 8% |
| BMI<sup>†</sup>, kg/m<sup>2</sup> | 27.5 (6.5) | 26.5 (6.7) | 28.1 (8.0) | 27.4 (7.8) | 29.9 (6.8) | 29.1 (7.2) |
| Current or former cigarette use | 47% | 47% | 44% | 44% | 85% | 85% |
| Diabetes mellitus | 3% | 3% | 6% | 6% | 5% | 5% |
| Hypertension | 46% | 46% | 52% | 52% | 48% | 48% |
| ASCVD Risk score<sup>†</sup>, % | 11.7 (13.3) | 7.5 (10.6) | 11.7 (11.7) | 9.2 (11.7) | 14.8 (14.0) | 10.5 (13.0) |
| Lipid panel<sup>‡</sup>, mg/dL |  |  |  |  |  |  |
| TC | 248 (48) | 203 (43) | 221 (52) | 190 (43) | 215 (60) | 177 (44) |
| LDL-C | 161 (40) | 120 (37) | 140 (46) | 110 (39) | 140 (53) | 104 (39) |
| HDL-C | 53 (19) | 55 (23) | 51 (20) | 55 (23) | 43 (15) | 45 (18) |
| TG | 139 (96) | 106 (75) | 127 (91) | 101 (73) | 145 (111) | 114 (88) |

Data presented as median (interquartile range) or percentage.
\*Statin users and nonusers were matched in a one user to two nonusers ratio by the following factors: age (within 3 years), sex, cigarette use, diabetes status, hypertension status
†ASCVD Risk score calculated using Pooled Cohort's Equation, which estimates 10-year percent risk of first ASCVD event(1).
‡Lipid panel and BMI based upon measurements on or before index date
AoURP, All Of Us Research Program; ASCVD, atherosclerotic cardiovascular disease; BMI, body mass index; GERA, Genetic Epidemiology Research on Adult Health and Aging; HDL-C, high-density lipoprotein cholesterol; LDL-C, low-density lipoprotein cholesterol; MVP, Million Veteran Program; NIH, National Institutes of Health; TC, total cholesterol; TG, triglycerides; VA, Veterans Affairs.

### Drug response phenotype

Among participants with no myocardial infarction at index, statin users had significantly reduced rates of the primary outcome in GERA (HR 0.77, 95% confidence interval [CI] 0.74-0.81; *P*=7.4E-26), AoURP (HR 0.86, 95% CI 0.75-0.98; *P*=.03), and MVP (HR 0.51, 95% CI 0.46-0.57; *P*=7.9E-35) compared to statin nonusers.

### Polygenic score

We observed strong correlations between the polygenic scores (i.e., participants with high polygenic risk from one score type tended to have high scores for the others). For example, within GERA (n=30,108), PRS2022 correlated with both 164SNP (Pearson’s r=0.43, *P*<1.0E-300) and metaGRS (r=0.62, *P*<1.0E-300). Additionally, metaGRS was also correlated with 164SNP (r=0.43, *P*<1.0E-300). As shown above, the correlations were strongest between PRS2022 and metaGRS. Supplementary Table 2 shows correlations between polygenic scores for the other cohorts.

### Coronary artery disease polygenic risk score and cardiovascular outcomes in statin nonusers

Higher polygenic score was associated with elevated risk of major adverse cardiovascular events. In GERA, this relationship was similar for PRS2022 (HR 1.24 per polygenic risk score standard deviation, 95% CI 1.21–1.28; *P*=9.4E-47), metaGRS (HR 1.18 per polygenic risk score standard deviation, 95% CI 1.15–1.21; *P*=1.3E-32), and 164SNP (HR 1.23 per polygenic risk score standard deviation, 95% CI 1.17–1.30; *P*=3.1E-15). Results for the full set of cohorts are available in Supplementary Tables 3 and 4.

### Coronary artery disease polygenic risk score and statin effectiveness

Among participants with no myocardial infarction at index, statin use was more effective for preventing the primary outcome compared to statin nonuse in the high polygenic score group versus the low score group across all cohorts for PRS2022 (interaction beta 0.19, standard error 0.07, interaction *P*=2.3E-3) and metaGRS (interaction beta 0.14, standard error 0.07, interaction *P*=0.02), but not 164SNP (interaction beta 0.09, standard error 0.07, interaction *P*=0.08). Figure 1 shows the comparison of HRs in GERA. Results were similar in the subset of participants with no ASCVD at index (Supplementary Table 5).

**Figure 1.**
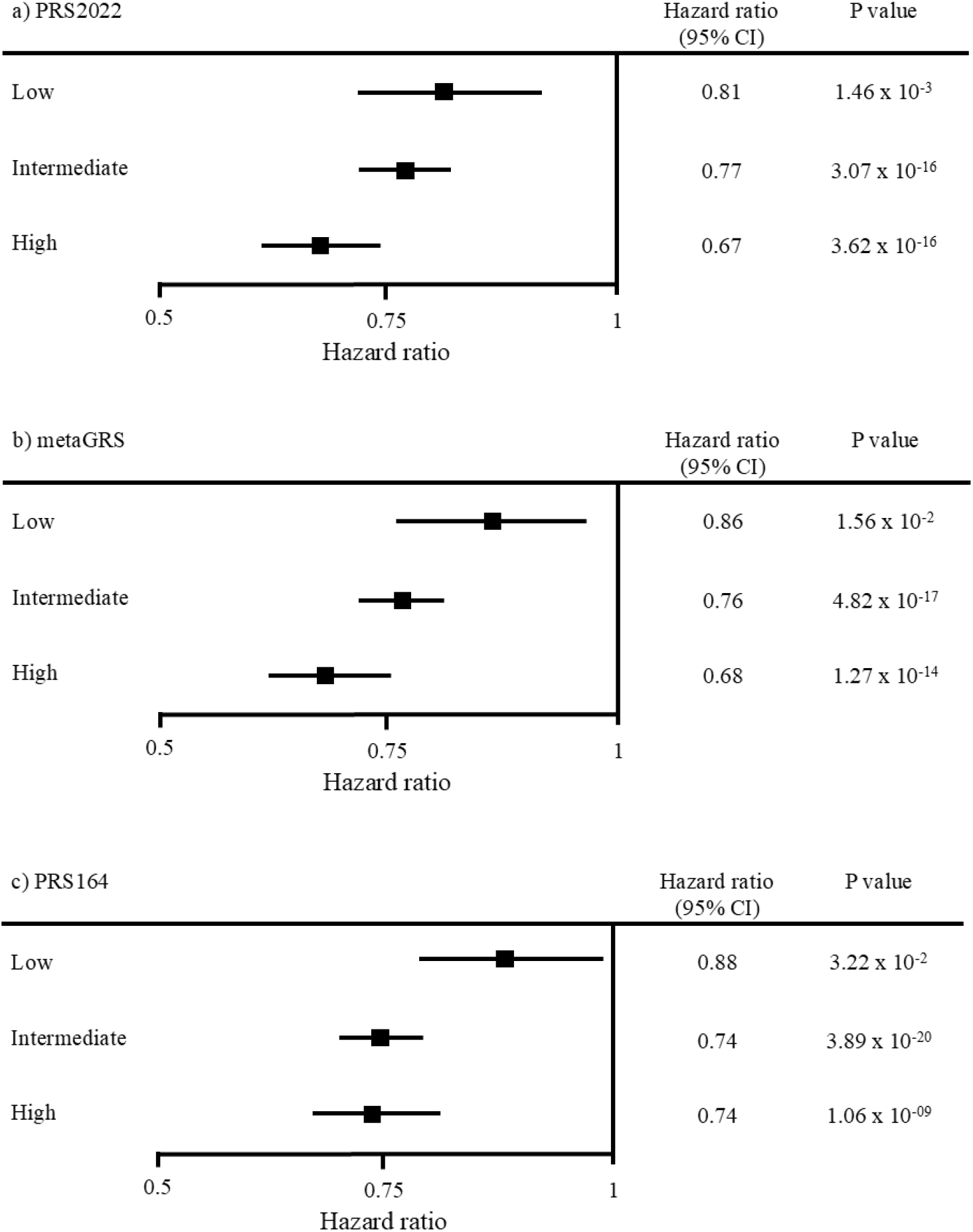
Statin effectiveness on incident major adverse cardiovascular events across coronary heart disease polygenic risk score groups for three different scoring types in GERA participants. Cox proportional hazard models of time-to-event were generated for statin users and matched nonusers among participants with no myocardial infarction at index to determine statin effectiveness in reducing incident major adverse cardiovascular events (defined as myocardial infarction, coronary revascularization, unstable angina, ischemic stroke, transient ischemia attack, or death). Covariates included age, sex, low-density lipoprotein cholesterol, diabetes, hypertension, and cigarette smoking. Across the various polygenic risk scoring types, increasing score was associated with enhanced statin effectiveness such that participants in the low polygenic risk score group (quintile 1) received the smallest benefit and participants in the high polygenic risk score group (quintile 5) experienced the largest benefit. Participants with polygenic risk scores in quintiles 2-4 made up the intermediate polygenic risk score group. Statin use was more effective in reducing major adverse cardiovascular events for the high polygenic score group versus the low score group with PRS2022 (interaction beta 0.19, standard error 0.08, interaction *P*=0.011), metaGRS (interaction beta 0.23, standard error 0.08, interaction *P*=0.001), and 164SNP (interaction beta 0.18, standard error 0.08, interaction *P*=0.009).

## Discussion

Our results provide clinical validation for the previously identified association between a coronary artery disease polygenic risk score and statin effectiveness. Despite markedly different clinical characteristics between multiple real-world cohorts, polygenic risk remained strongly associated with relative risk reduction of cardiovascular outcomes from statins among patients remaining adherent to therapy. This is the first investigation of statin relative risk reduction using polygenic scores that include variants below thresholds of genome-wide significance.

Over the past decade, numerous polygenic risk scores derived from genome-wide association studies of coronary artery disease have demonstrated a statistically significant association with cardiovascular outcomes.^11,12^ Subsequent studies have shown that these scores may be useful in reclassifying patients into new baseline coronary artery disease risk categories.^13,14^ In contrast, other studies have shown conflicting evidence for the performance of polygenic risk scores in improving baseline ASCVD risk stratification, casting doubt on their potential clinical translation.^15^

A key limitation for all these initial studies is that they assume statin-relative risk reduction is uniform across the population. However, in addition to modifying baseline coronary artery disease risk, these polygenic risk scores were found to predict efficacy of statin treatment in preventing coronary artery disease. In particular, evidence from genetic substudies of five statin trials showed that high coronary artery disease polygenic risk predicts a greater magnitude of statin relative risk reduction.^3,4^ Importantly, these associations were independent of established clinical and demographic risk factors of coronary artery disease, including family history, a factor often considered to be a proxy for genetic information. Furthermore, the association was independent of statin-induced LDL-C lowering, challenging the findings of the Cholesterol Treatment Trialists’ Collaborators that the magnitude of statin relative risk reduction is only related to statin-induced LDL-C lowering.^2^ Altogether, these results suggest that a pharmacogenomics approach to the implementation of coronary artery disease polygenic risk scores (not only to reclassify baseline coronary artery disease risk, but also for prescribing therapy) may enhance clinical decision-making for statin therapy assignment.

We recently demonstrated that the ability of a coronary artery disease polygenic risk score to predict statin relative risk reduction, originally reported in randomized controlled trials, extends to real-world participants undergoing routine care.^5^ In particular, those participants with the highest polygenic risk had a stronger benefit especially compared to those with the lowest. The current study sought to address a variety of hypotheses generated from the findings of the previous study in a larger study population with more extensive statin cardiovascular benefit phenotyping.

The enhanced phenotyping was a key strength of the current study. Compared to prior studies, we were better able to characterize cardiovascular status both at index and at the time of an event. For example, our major adverse cardiovascular event outcome included coronary artery revascularization procedures as well as unstable angina. Furthermore, we were able to generate an ASCVD-free primary prevention cohort. This cohort is more stringent than the previous myocardial infarction-free study population because it considers stable angina, abdominal aortic aneurysm, and other cardiovascular diseases that are often overlooked when determining eligibility for primary prevention studies. More robust and detailed phenotypes allowed for our results to better align with statin benefit groups, improving potential translation of findings into clinical care.

We were ultimately able to validate the association between coronary artery disease polygenic risk and statin benefit in this more robust primary prevention cohort. Interestingly, we found that both PRS2022 and metaGRS coronary artery disease polygenic scores were more strongly associated with statin relative risk reduction compared to 164SNP. Whereas 164SNP includes only variants that have been previously associated with coronary artery disease at the level of genome-wide significance, PRS2022 and metaGRS were generated using more advance techniques. In particular, PRS2022 was derived using LDPred.^8^ In contrast, metaGRS is from a meta-analysis of multiple coronary artery disease polygenic scores. Each score within metaGRS uses either a variation of linkage disequilibrium pruning or false discovery rate.^7^ Consequently, the genetic variant counts for PRS2022 (2.3 million variants) and metaGRS (1.7 million variants) far surpassed that of 164SNP (164 variants). It has been established that the inclusion of subthreshold variants in coronary artery disease polygenic scores provides improved prediction performance.^11^ The current study extends this finding by applying that principle to statin relative risk reduction for cardiovascular outcomes.

A limitation of the current study is that confounding can never be completely ruled out with observational data. As such, the associations reported must be interpreted with caution. However, the use of multiple independent cohorts strengthens the robustness and generalizability of our findings, which may mitigate any potential confounding unique to any single dataset. Future studies, including the investigation of sub-threshold-inclusive polygenic risk scores in randomized controlled trial genetic substudies, would help reinforce the current results.

In summary, we demonstrated that the association between coronary artery disease polygenic risk scores and statin relative risk reduction can be enhanced with the inclusion of both genome-wide significant and subthreshold variants. These results help to pave the way for future investigation regarding the clinical utility of candidate statin polygenic scores as precision medicine tools. Crucially, additional studies are necessary to identify polygenic scores that can be applied to other statin benefit groups (e.g., secondary prevention) and broader populations.

## Data Availability

All summary-level data produced in the present study are available upon reasonable request to the authors

## Acknowledgments

We thank the Kaiser Permanente Northern California members who have generously agreed to participate in the Kaiser Permanente Genetic Epidemiology Research on Adult Health and Aging (GERA) cohort. The development of GERA was supported by grants from the Robert Wood Johnson Foundation, the Wayne and Gladys Valley Foundation, the Ellison Medical Foundation, and Kaiser Permanente Community Benefit Programs. This work was supported using resources and facilities of the VA Informatics and Computing Infrastructure (VINCI), including data analytics conducted by its Precision Medicine research team, which is funded under the research priority to Put VA Data to Work for Veterans (VA ORD 24-D4V-02). This research is also supported by Million Veteran Program award MVP003/028 I01-BX003362. Kathryn Pridgen assisted with manuscript preparation. This article does not reflect the position or policy of the Department of Veterans Affairs or the United States government.

