## Supplement for "Clinical validation of a statin-benefit polygenic score using real-world cohorts of primary prevention participants"

**Supplemental Methods**

Data source (continued)

The National Institutes of Health All of Us research program (AoURP) is a public EHR-linked biobank with a goal of >1,000,000 enrollees 18 years and older.^1^ We used the All of Us Controlled Tier Dataset v7 (curated version 2022Q4R9), which was the most recent data version at the time of data analysis and included data from participants who enrolled between 2018 and 2021. Among >600,000 enrolled participants enrolled at the time, 60% identified as woman and 39% identified as man. The proportion of participants aged 18-49, 50-69, and 70+ at enrollment were 39%, 40%, and 21%, respectively. For the current investigation, there were ~245,394 participants with adequate EHR, survey, and genomic data from which a study population was selected. The EHR included pharmacy records, diagnostic/procedure codes, vital records, and laboratory measurements. Genetic polymorphism data was derived from the AoURP Allele Count/Allele Frequency (ACAF) reduced dataset.

Kaiser Permanente Northern California (KPNC) region medical care plan is an integrated health care delivery system with a population of >3.3 million people. We used the Virtual Data Warehouse from the Division of Research under KPNC as a source for the health records used in the current study. KPNC EHR data included pharmacy records, diagnostic/procedure codes, vital records, and laboratory measurements between 1995 and 2022. The Genetic Epidemiology Research on Adult Health and Aging (GERA) cohort is a biobank linked to the KPNC EHR and contains 110,266 participants with genome-wide genotyping data. Genotyping data was generated from one of four Affymetrix Axiom arrays based on self-identified race/ethnicity.^2^ Imputation was performed to the 1000 Genomes Project phase 1 as previously described.^3^ In addition to EHR and genomic data, participants also completed self-administered surveys for sociodemographic information. Most participants self-identify as non-Hispanic White (81%). We used both survey and genomic data from GERA for the current investigation.

The Veterans Health Administration (VHA) is a large, integrated health system serving >9 million Veterans across the United States.^4^ We used the Corporate Data Warehouse, the EHR of the VHA, which includes patient demographics, smoking status, body mass index, pharmacy records, diagnostic/procedure codes, vital records, and laboratory measurements. Data spanned 2000 to 2023 (with required statin initiation between 2000 and 2021 for those who met the criteria of a statin user). Million Veteran Program (MVP) is the biobank linked to the VHA EHR and includes ~900,000 participants with genome-wide genotyping data. Most participants self-identified as non-Hispanic White (71%), with 18% identifying as non-Hispanic Black and 8% as Hispanic. Genotyping data was generated from a customized Affymetrix Axiom array before undergoing quality control. Imputation was done with the 1000 Genomes phase 3 v5 reference panel using EAGLE v2.4 and Minimac4.^5^

Drug response phenotype (continued)

For the Pooled Cohort Equations, we considered the following therapies to define a antihypertensive user: 'acebutolol', 'aliskiren', 'amiloride', 'amlodipine', 'atenolol', 'benazepril', 'bisoprolol', 'candesartan', 'captopril', 'carvedilol', 'chlorothiazide', 'chlorthalidone', 'clonidine', 'diltiazem', 'doxazosin', 'enalapril', 'eplerenone', 'felodipine', 'fosinopril', 'guanabenz', 'hydralazine', 'hydrochlorothiazide', 'indapamide', 'irbesartan', 'isosorbide', 'isradipine', 'labetalol', 'lisinopril', 'losartan', 'methyldopa', 'metoprolol', 'minoxidil', 'moexipril', 'nadolol', 'nebivolol', 'nicardipine', 'nifedipine', 'nimodipine', 'nisoldipine', 'olmesartan', 'penbutolol', 'perindopril', 'pindolol', 'prazosin', 'propranolol', 'quinapril', 'ramipril', 'reserpine', 'sotalol', 'spironolactone', 'telmisartan', 'terazosin', 'trandolapril', 'triamterene', 'valsartan', 'verapamil'.

Supplementary Table 1.

Diagnostic and procedural codes used for each cohort ^a^

| **KPNC GERA** | **NIH AoURP** | **VA MVP** |
| --- | --- | --- |
| No ASCVD primary prevention cohort   1. MI (ICD-9: 410; ICD-10: I21, I22, I23) 2. Old MI (ICD-9: 412; ICD-10: I25.2) 3. Stable angina (ICD-9: 413, 414; ICD-10: I20.1, I20.8, I20.9, I24.8, I24.9, I25.1, I25.8, I25.9) 4. Unstable angina (ICD-9: 411; ICD-10: I20.0, I24.0) 5. Revascularization    a. CABG (ICD-9: V45.81; CPT: 33510-33536)    b. PCI (ICD-9: V45.82; ICD-10: Z95.5; CPT: 92920–92924, 92928, 92929, 92933, 92934, 92937, 92938, 92941, 92943, 92944)    c. Angioplasty (ICD-9: Z98.61; CPT: 92980-92982, 92984, 92995, 92996)    d. Endovascular (CPT: 37220, 37221, 37222, 37223, 37224, 37226, 37228)    e. Stent (CPT: C1874, C1875, C1876, C1877) 6. Ischemic stroke/carotid stenosis (ICD-9: 433, 434, 436, 438; ICD-10: I63, I65, I66) 7. TIA (ICD-9: 435; ICD-10: G45) 8. PAD (ICD-9: 440.20–24, 440.0 [not “440”], 440.4, 443.9; ICD-10: I70) 9. Abdominal aortic aneurysm (ICD-9: 441; ICD-10: I71)   Note: an ICD-9 code for old MI (412) during follow-up without any MI codes prior to that was indicative of “MI before index” | Same as 1st column | No ASCVD primary prevention cohort   1. MI (ICD-9: 410; ICD-10: I21, I22, I23) 2. Old MI (ICD-9: 412; ICD-10: I25.2) 3. Stable angina (ICD-9: 413, 414; ICD-10: I20.1, I20.8, I20.9, I24.8, I24.9, I25.1, I25.8, I25.9) 4. Unstable angina (ICD-9: 411; ICD-10: I20.0, I24.0) 5. Revascularization    a. CABG (ICD-9: V45.81; CPT: 33510-33536)    b. PCI (ICD-9: V45.82; ICD-10: Z95.5; CPT: 92920–92924, 92928, 92929, 92933, 92934, 92937, 92938, 92941, 92943)    c. Angioplasty (ICD-10: Z98.61; CPT: 92980-92982, 92984, 92995, 92996)    d. Endovascular (CPT: 37220, 37221, 37222, 37223, 37224, 37226, 37228)    e. Stent (CPT: C1874, C1875, C1876, C1877)    f. Other revascularization (ICD 9-PCS: 0.66, 36.00, 36.01, 36.02, 36.03, 36.04, 36.05, 36.06, 36.07, 36.09, 36.10, 36.11, 36.12, 36.13, 36.14, 36.15, 36.16, 36.17, 36.19, and 36.2; ICD 10-PCS: 021008W, , 021009C, 021009F, 021009W, 02100A3, 02100A8, 02100A9, 02100AC, 02100AW, 02100J3, 02100J8, 02100J9, 02100JW, 02100K3, 02100K8, 02100K9, 02100KW, 02100Z3, 02100Z8, 02100Z9, 02100ZC, 02103D4, 021049W, 02104A8, 02104A9, 02104AW, 02104D4, 02104K9, 02104KW, 02104Z3, 02104Z9, 021109C, 021109F, 021109W, 02110A3, 02110A8, 02110A9, 02110AC, 02110AW, 02110J9, 02110JW, 02110KW, 02110Z3, 02110Z8, 02110Z9, 02110ZC, 02110ZF, 021149W, 02114J9, 02114Z9, 021209C, 021209W, 02120A3, 02120A9, 02120AC, 02120AW, 02120J3, 02120J9, 02120JW, 02120K3, 02120K9, 02120KW, 02120Z3, 02120Z8, 02120Z9, 02123D4, 021249W, 02124D4, 02124Z3, 021309W, 02130A8, 02130A9, 02130AW, 02130J3, 02130J8, 02130J9, 02130JW, 02130KW, 02130Z9, 02134D4, 02134KW, 027004Z, 027005Z, 027006Z, 02700D6, 02700DZ, 02700Z6, 02700ZZ, 027034Z, 027035Z, 027036Z, 027037Z, 02703D6, 02703DZ, 02703EZ, 02703FZ, 02703GZ, 02703TZ, 02703Z6, 02703ZZ, 027044Z, 027045Z, 027046Z, 027047Z, 02704D6, 02704DZ, 02704EZ, 02704FZ, 02704GZ, 02704Z6, 02704ZZ, 027104Z, 027105Z, 02710DZ, 02710ZZ, 027134Z, 027135Z, 027136Z, 027137Z, 02713D6, 02713DZ, 02713EZ, 02713F6, 02713FZ, 02713GZ, 02713Z6, 02713ZZ, 027144Z, 027145Z, 027146Z, 027147Z, 02714DZ, 02714EZ, 02714GZ, 02714ZZ, 027205Z, 02720FZ, 027234Z, 027235Z, 027236Z, 027237Z, 02723D6, 02723DZ, 02723EZ, 02723F6, 02723FZ, 02723G6, 02723GZ, 02723Z6, 02723ZZ, 027244Z, 027245Z, 027246Z, 02724DZ, 02724EZ, 02724FZ, 02724ZZ, 027304Z, 02730DZ, 02730ZZ, 027334Z, 027335Z, 027336Z, 027337Z, 02733DZ, 02733G6, 02733GZ, 02733ZZ, 027344Z, 027345Z, 027347Z, 02734DZ, 02734GZ, 02734ZZ, 02C00Z6, 02C00ZZ, 02C03Z6, 02C03ZZ, 02C04ZZ, 02C10ZZ, 02C13Z6, 02C13ZZ, 02C14ZZ, 02C33ZZ, and 02C43ZZ) 6. Ischemic stroke/carotid stenosis (ICD-9: 433, 434, 436, 438; ICD-10: I63, I65, I66) 7. TIA (ICD-9: 435; ICD-10: G45) 8. PAD (ICD-9: 440.20–24, 440.0 [not “440”], 440.4, 443.9; ICD-10: I70) 9. Abdominal aortic aneurysm (ICD-9: 441; ICD-10: I71)   Note: an ICD-9 code for old MI (412) during follow-up without any MI codes prior to that was indicative of “MI before index” |
| MACE   1. Myocardial infarction    a. ICD-9: 410; ICD-10: I21, I22    b. ICD-9: 411; ICD-10: I20.0 plus elevated troponin (>10ng/ml) within 7 days of the code    c. Participants with ICD-9: 411; ICD-10: I20.0 plus uncertain or missing troponin levels were excluded from the study population 2. Revascularization    a. CABG (CPT: 33510-33536)    b. PCI (CPT: 92920–92924, 92928, 92929, 92933, 92934, 92937, 92938, 92941, 92943, and 92944)    c. Angioplasty (CPT: 92980-92982, 92984, 92995, 92996) 3. Unstable angina (ICD-9: 411; ICD-10: I20.0) 4. Ischemic stroke (ICD-9: 433.x1, 434.x1; ICD-10: I63) 5. TIA (ICD-9: 435; ICD-10: G45) 6. Death from any cardiovascular cause (ICD-9: 390–459; ICD-10: I*)    a. The position does not matter (can be primary cause of death, secondary, etc.) | MACE   1. Myocardial infarction    a. ICD-9: 410; ICD-10: I21, I22    b. ICD-9: 411; ICD-10: I20.0 plus elevated troponin (>10ng/ml) within 7 days of the code    c. Participants with ICD-9: 411; ICD-10: I20.0 plus uncertain or missing troponin levels were excluded from the study population 2. Revascularization    a. CABG (CPT: 33510-33536)    b. PCI (CPT: 92920–92924, 92928, 92929, 92933, 92934, 92937, 92938, 92941, 92943, and 92944)    c. Angioplasty (CPT: 92980-92982, 92984, 92995, 92996) 3. Unstable angina (ICD-9: 411; ICD-10: I20.0) 4. Ischemic stroke (ICD-9: 433.x1, 434.x1; ICD-10: I63) 5. TIA (ICD-9: 435; ICD-10: G45) 6. Death from any cause | MACE   1. Same as 2nd column |
| Hypertension   1. Two or more diagnosis codes (ICD-9: 401; ICD-10: I10) 2. The first code is considered the date of diagnosis | Same as 1st column | Same as 1st column |
| Diabetes   1. Definition    a. ≥1 T2D ICD codes PLUS ≥1 T2D drug fills    b. ≥1 T2D ICD codes PLUS ≥2 T2D/T1D drug fills with the T2D fill occurring before the earliest T1D drug fill    c. ≥2 T2D ICD codes PLUS ≥1 T1D drug fills    d. ≥1 T2D ICD codes PLUS ≥1 positive lab results    e. ≥1 T1D ICD code automatically precludes a T2D case 2. T2D ICD codes (ICD-9: 250.x0, 250.x2; ICD-10: E11) 3. T1D ICD codes (ICD-9: 250.x1, 250.x3; ICD-10: E10) 4. T2D drugs ('acetohexamide', 'tolazamide', 'chlorpropamide', 'glipizide', 'glyburide', 'glimepiride', 'repaglinide', 'nateglinide', 'metformin', 'rosiglitazone', 'pioglitazone', 'troglitazone', 'acarbose', 'miglitol', 'sitagliptin', 'exenatide', 'saxagliptin', 'linagliptin', 'liraglutide', 'semaglutide', 'canagliflozin', 'dapagliflozin', 'empagliflozin', 'alogliptin', 'colesevelam', 'albiglutide', 'dulaglutide', 'lixisenatide', 'ertugliflozin', 'tolbutamide') 5. T1D drugs (insulin', 'pramlintide', 'insulin glulisine', 'insulin lispro', 'insulin aspart', 'insulin glargine', 'insulin detemir', 'insulin degludec', 'insulin NPH') 6. Positive lab results (A1C ≥6.5%; fasting glucose ≥126 mg/dl, random glucose ≥200) | Same as 1st column | Same as 1st column |

^a^ ICD codes without a decimal point are inclusive of all subcodes within (e.g., ICD-10 “I22” includes I22.0, I22.1, I22.2, I22.8, I22.9, etc.; ICD-10 “I20.0” does not include I20.1, I20.2, I20.8, I20.9, etc.) whereas if an “x” or “*” implies that any number can go into that position (e.g., ICD-9 “433.x1” includes 433.01, 433.11, 433.21, 433.31, etc.; ICD-9 “433.x1” does not include 433.00, 433.10, 433.20, etc.)

A1C, hemoglobin A1C; ASCVD, atherosclerotic cardiovascular disease; CABG, coronary artery bypass graft; CPT, Current Procedural Terminology; dl, deciliter; GERA, Genetic Epidemiology Research on Aging; ICD, International Classification of Diseases; KPNC, Kaiser Permanente Northern California; MACE, major adverse cardiovascular events; MI, myocardial infarction; ml, milliliter; MVP, Million Veteran Program; ng, nanograms; PAD, peripheral arterial disease; PCI, percutaneous coronary intervention; T1D, type 1 diabetes; T2D, type 2 diabetes; TIA, transient ischemic attack; VA, Veterans Affairs.

Supplementary Table 2.

Correlations between polygenic risk scores. ^a^

|  | **GERA** | | | **NIH AoURP** | | | **VA MVP** | | |
| --- | --- | --- | --- | --- | --- | --- | --- | --- | --- |
|  | PRS2022 | metaGRS | 164SNP | PRS2022 | metaGRS | 164SNP | PRS2022 | metaGRS | 164SNP |
| PRS2022^6^ |  | 0.62* | 0.43* |  | 0.64* | 0.37* |  | 0.62* | 0.00 |
| metaGRS^7^ |  |  | 0.43* |  |  | 0.37* |  |  | 0.00 |
| 164SNP^8^ |  |  |  |  |  |  |  |  |  |

^a^ Peason’s r; * indicates *P*< 0.001

Supplementary Table 3.

Risk of incident major adverse cardiovascular events per polygenic risk score standard deviation in statin nonusers with no myocardial infarction history at index

|  | **GERA** | | **NIH AoURP** | | **VA MVP** | |
| --- | --- | --- | --- | --- | --- | --- |
|  | HR (95% CI) | P-value | HR (95% CI) | P-value | HR (95% CI) | P-value |
| PRS2022^6^ | 1.24 (1.21—1.28) | 9.4E-47 | 1.26 (1.16—1.36) | 3.8E-9 | 1.13 (1.08—1.17) | 6.4E-9 |
| metaGRS^7^ | 1.18 (1.15—1.21) | 1.3E-32 | 1.23 (1.14—1.32) | 1.2E-7 | 1.07 (1.03—1.11) | 1.3E-3 |
| 164SNP^8^ | 1.23 (1.17—1.30) | 3.1E-15 | 1.03 (0.96—1.11) | 0.43 | 1.00 (0.97—1.05) | 0.80 |

AoURP, All Of Us Research Program; CI, confidence interval; GERA, Genetic Epidemiology Research on Adult Health and Aging; HR, hazard ratio; MVP, Million Veteran Program; NIH, National Institutes of Health; VA, Veterans Affairs.

Supplementary Table 4.

Risk of incident major adverse cardiovascular events per polygenic risk score standard deviation in statin nonusers with no atherosclerotic cardiovascular disease at index

|  | **GERA** | | **NIH AoURP** | | **VA MVP** | |
| --- | --- | --- | --- | --- | --- | --- |
|  | HR (95% CI) | P-value | HR (95% CI) | P-value | HR (95% CI) | P-value |
| PRS2022^6^ | 1.23 (1.19—1.27) | 2.9E-37 | 1.27 (1.16—1.39) | 1.1E-7 | 1.12 (1.07—1.17) | 4.3E-7 |
| metaGRS^7^ | 1.17 (1.14—1.21) | 1.1E-26 | 1.24 (1.13—1.35) | 2.4E-6 | 1.08 (1.04—1.13) | 5.2E-4 |
| 164SNP^8^ | 1.22 (1.15—1.29) | 2.5E-12 | 1.00 (0.92—1.09) | 0.97 | 1.00 (0.96—1.05) | 0.85 |

AoURP, All Of Us Research Program; CI, confidence interval; GERA, Genetic Epidemiology Research on Adult Health and Aging; HR, hazard ratio; MVP, Million Veteran Program; NIH, National Institutes of Health; VA, Veterans Affairs.

Supplementary Table 5.

Interaction of statin effectiveness on major adverse cardiovascular events in low polygenic versus high polygenic risk score groups ^a^ among participants with no atherosclerotic cardiovascular disease at index

|  | Interaction beta ^b^ | Standard error | P-value |
| --- | --- | --- | --- |
| PRS2022^6^ | 0.12 | 0.07 | 0.055 |
| metaGRS^7^ | 0.18 | 0.07 | 0.006 |
| 164SNP^8^ | 0.09 | 0.07 | 0.116 |

^a^ Meta-analysis across participants from GERA, NIH AoURP, VA MVP cohorts for each risk score. Low and high are based on the bottom and top 20% of participants of polygenic risk for each score, respectively.

^b^ Beta is the difference in hazard ratios between polygenic risk score groups as follow: low subtracted by high. Thus, a positive interaction beta indicates that statin is more effective in high polygenic risk score group versus low.

AoURP, All Of Us Research Program; GERA, Genetic Epidemiology Research on Adult Health and Aging; MVP, Million Veteran Program; NIH, National Institutes of Health; VA, Veterans Affairs.
